# Outdoor Physical Activity as a Confounder for Vitamin D Status: A Scoping Review Protocol

**DOI:** 10.1101/2023.01.30.23285088

**Authors:** Patrick G. Corr, Alia M. Badawi, Stephanie H. Vu, Leigh A. Frame

**Author notes:** Corresponding Author, (PGC). These authors contributed equally to this work and have approved the following manuscript (Patrick G. Corr, Alia M. Badawi, Stephanie H. Vu). This author contributed equally to this work and served as senior author.

## Abstract

**Background:** Vitamin D (vitD) has been correlated with a number of health outcomes and disease states; however, these relationships are likely confounded by the role of outdoor physical activity (PA). Currently, there are few studies that consider confounders to vitD status and the potentially spurious relationships between PA and vitD status.

**Methods:** This scoping review will employ Arksey and O’Malley’s five-stage approach. Publications will be identified through five databases (CINAHL, Cochrane Library, PubMed, Scopus, and SportDISCUS) and reviewed in Covidence.

**Results:** The findings are expected to help identify what information is presently available regarding the correlation between outdoor PA and vitD status. The results of this scoping review will help to inform a quantitative analysis of the National Institutes of Health (NIH) All of Us database.

**Discussion:** It is unclear what effect outdoor PA may have on the relationship between vitD status and health outcomes and if any portion of this relationship is spurious. This scoping review will initiate an exploration of the relationship between vitD and PA to determine the strength and magnitude of potential confounders. Ultimately, we seek to reassess previous findings and determine the true effect of vitD alone by isolating the effect of outdoor PA.

## Introduction

Vitamin D (vitD) is a well-known fat-soluble hormone that is often discussed in reference to health benefits and disease states. Most commonly, it is known for its ability to facilitate calcium and phosphorus absorption in the body—the function that earned it distinction as a vitamin. More recently, a role for vitD in immune function has been elucidated. VitD is primarily obtained through endogenous production of 25-hydroxyvitamin D_3_ (cholecalciferol) from 7-dehydrocholesterol (protovitamin D_3_) after skin is exposed to sunlight (photoproduction); this process subsequently triggers 7-dehydrocholesterol (7-DHC) in the epidermis to be converted to provitamin D_3_, which then spontaneously isomerizes to vitamin D_3_ [2]. Photoproduction is the primary source of vitD for humans. Smaller amounts of vitD are also obtained exogenously through the diet as vitD_3_ and vitD_2_ (ergocalciferol); though, dietary vitD is a minor contributor to overall vitD status [2]. Additionally, vitD_3_ is more effective at raising overall serum 25(OH)D concentration than dietary or supplemental vitD_2_, even when taken at similar doses [3]. Both forms require activation through hydroxylation reactions that take place in the liver and in the kidneys to exert physiologic effects. These hydroxylation steps produce vitD metabolites: the circulating form 25-hydroxyvitamin D [25(OH)D] and the activated form 1,25-dihydroxyvitamin D [1,25(OH)2D] [4]. Serum 25(OH)D_3_, 25(OH)D_2_, and total 25(OH)D are used to quantify vitD status [5]. Clinically, total 25(OH)D alone is typically used, overlooking the important physiological differences between vitD_2_ and vitD_3_ [5].

### Background and Rationale

Although dietary vitamin D (vitD), which is predominantly vitD_3_, has been identified as a minor contributor to overall vitD status, the photoproduction of vitD_3_ during exposure to ultraviolet B (UV-B) radiation has been well-researched and identified as the primary source in humans. Therefore, time spent outdoors may be used as a proxy, convenience measure for vitD photoproduction. Hence, outdoor physical activity (PA) in existing vitD-related research is utilized as a representation of UV-B exposure; however, the relationship between outdoor PA and vitD status both dependent upon and independent of UV-B exposure and potential confounding effects is not well studied [2,3,5,7].Health benefits from outdoor PA that are not at all related to vitD status (i.e. independent relationships) as well as benefits that are directly related to vitD (i.e. dependent relationships) are well established in the literature. The independent benefits of outdoor PA may lead researchers to inaccurately report spurious associations between these two variables. Thus, the potential for these spurious relationships may throw off exercise science and nutrition research as it relates to vitD status and related health outcomes.

The strength and magnitude of the confounding between vitD and PA has not been clearly described to our knowledge. This is despite research on vitD and health outcomes seeing substantial growth, especially with the 2011 report on dietary reference intakes for vitD from the then Institute of Medicine, now the National Academy of Medicine (**Error! Reference source not found**.) [8]. Whether these publications are potentially biased by or include spurious associations is unclear; however, the potential for such spurious associations to alter the results or at least the magnitude of those results is great. Therefore, the potential for spurious relationship related to vitD and PA must be explored in depth. This scoping review seeks to identify where and how these relationships are discussed currently in the literature as an initial step.

**Figure 1:**
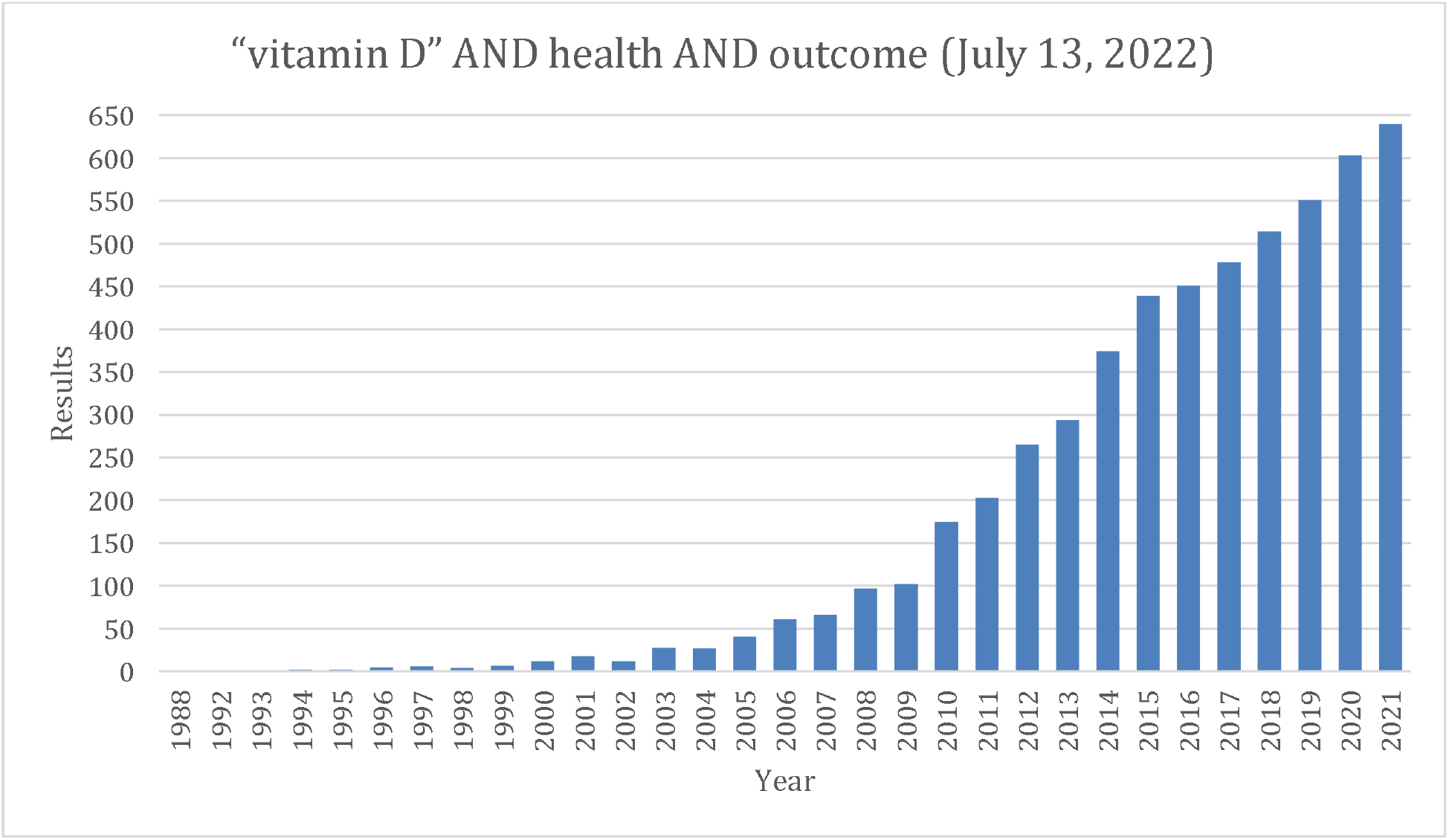
PubMed.gov Search Results

### Objectives

This scoping review is the first phase in a comprehensive study considering the strength of correlation between outdoor PA and vitD status, potential confounding effects, and previously identified spurious relationships between outdoor PA and vitD. Following the completion of this scoping review, a quantitative study will be conducted through the National Institutes of Health (NIH) All of Us Research Database analyzing variables of interest including 25(OH)D_3_, 25(OHD)D_3_ + D_2_, total 25(OH)D, calciferol (vitD_2_), cholecalciferol (vitD_3_), and FitBit data [5, 9]. The All of Us Research Database contains several markers of vitD status and includes the ability to distinguish vitD_3_, which is from endogenous photoproduction, dietary sources, and dietary supplements, and vitD_2_, which is driven predominantly by prescription supplementation and has a lower affinity for the vitD receptor [3, 5].

### Timeline

This scoping review was initiated in August 2022, reviewed by members of the research team, and shared with university reference librarians for initial feedback. The search strategy was finalized in October 2022 and initial review of the titles and abstracts was launched. Data extraction and analysis will begin in January 2023 and continue through February 2023. The results of this scoping review will be summarized and reported in March 2023.

### Methods & Protocol Design

This scoping review is designed to follow the five-stage process as described by Arksey and O’Malley [1]:

1. Identifying the Research Question

2. Identifying Relevant Studies

3. Study Selection

4. Charting the Data

5. Collecting, Summarizing and Report the Data

These distinct steps are discussed in greater detail below and provide a roadmap for the scoping review protocol we present.

### Stage One: Identifying the Research Question

Prior to developing the scoping review protocol, the research team conducted an initial, limited review of research considering the role of endogenous and exogenous vitD and their influence on overall vitD status as well as the correlations between vitD and various health outcomes and disease states. Through this initial research, a number of confounders to vitD status were identified; the largest identified gap was the relationship between outdoor PA and vitD status. With a clear background of the issue and the primary objective of this study in mind, the research team identified the following research questions to guide this scoping review:

1. What correlations exist between outdoor PA and vitD status?
2. How might regular outdoor PA improve health outcomes and/or disease conditions independently of vitD, as opposed to dependent upon vitD?
  a. What spurious relationships between vitD and outdoor PA have been previously identified?
3. What is the potential for and magnitude of confounding via a spurious relationship?

### Stage Two: Identification of Relevant Studies

The second stage of Arksey and O’Malley’s process for developing effective scoping reviews calls for researchers to clearly delineate how they will select relevant studies [1]. The literature identified through this scoping review will be housed in Covidence, a tool to facilitate and organize review and analysis. We provide a detailed overview of our search strategy, inclusion and exclusion criteria, and our process for reviewing literature below.

### Search Strategy & Information Sources

The search strategy for this scoping review was developed in consultation with a reference librarian at the George Washington University’s Himmelfarb Health Sciences Library to ensure a comprehensive approach to collecting data. The review will include published manuscripts retrievable from the electronic databases through CINAHL, the Cochrane Database of Systematic Reviews, PubMed, Scopus, and SportDISCUS. A detailed overview of the search terms for each of these databases is provide in Table 1.

**Table 1:**
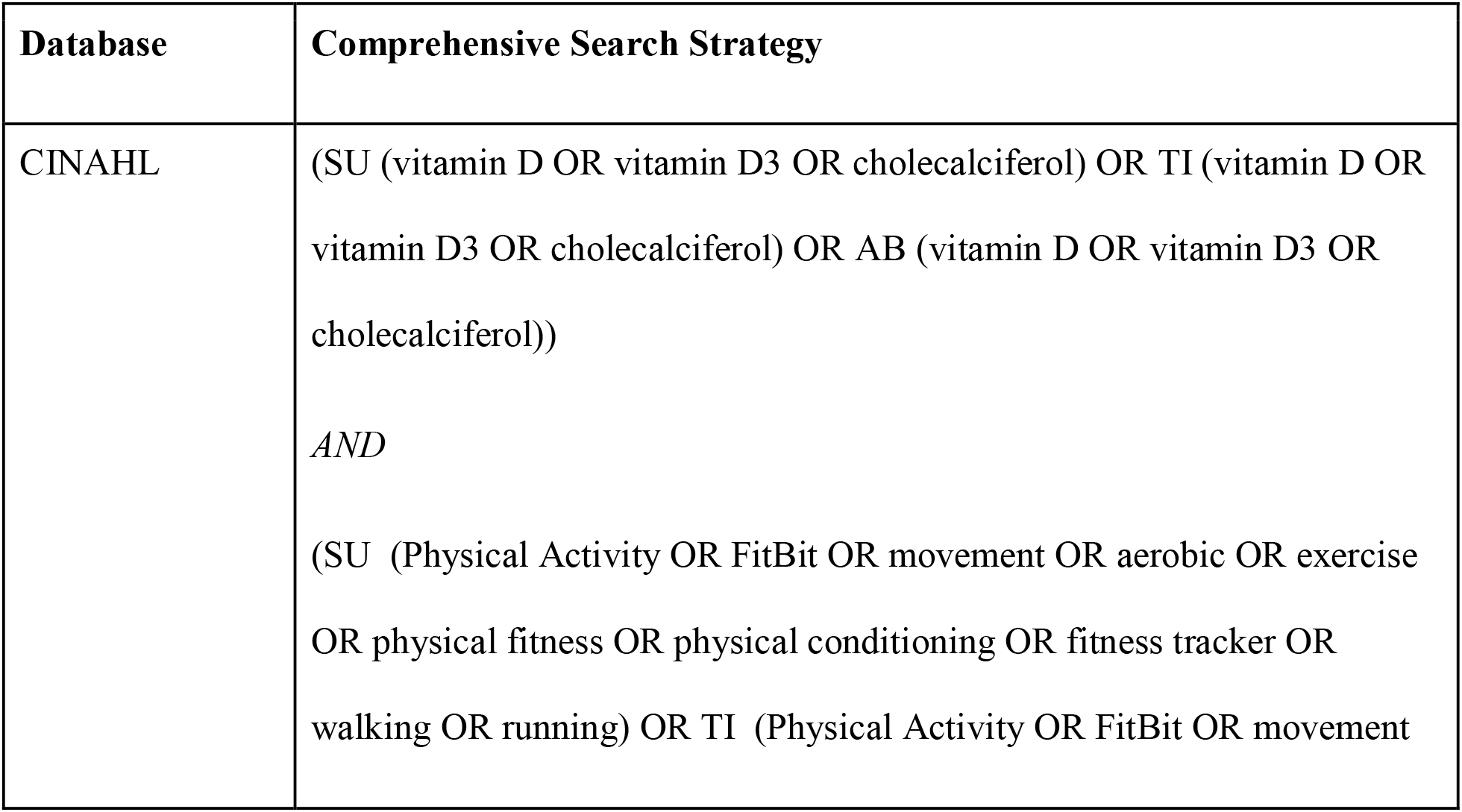
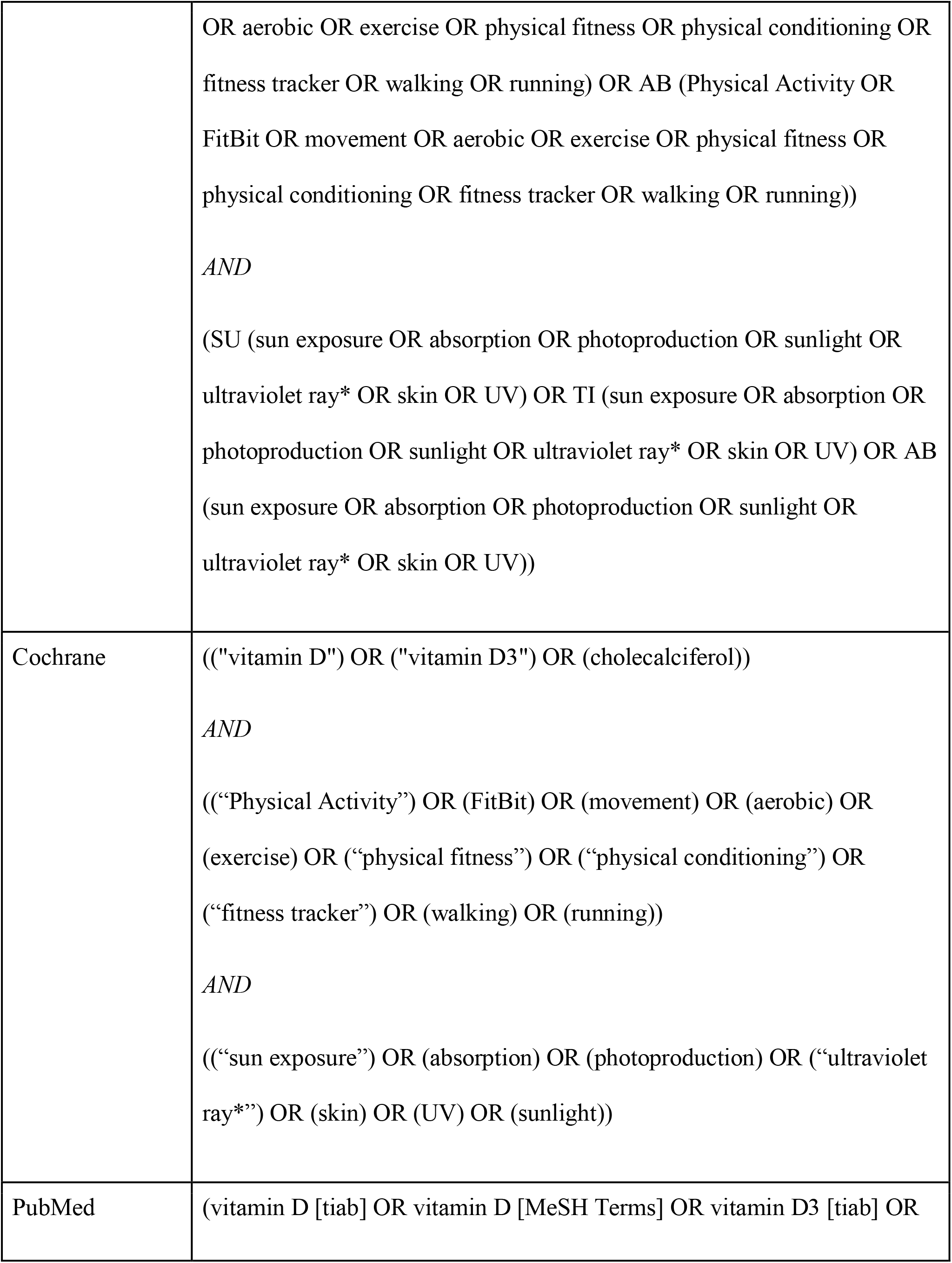
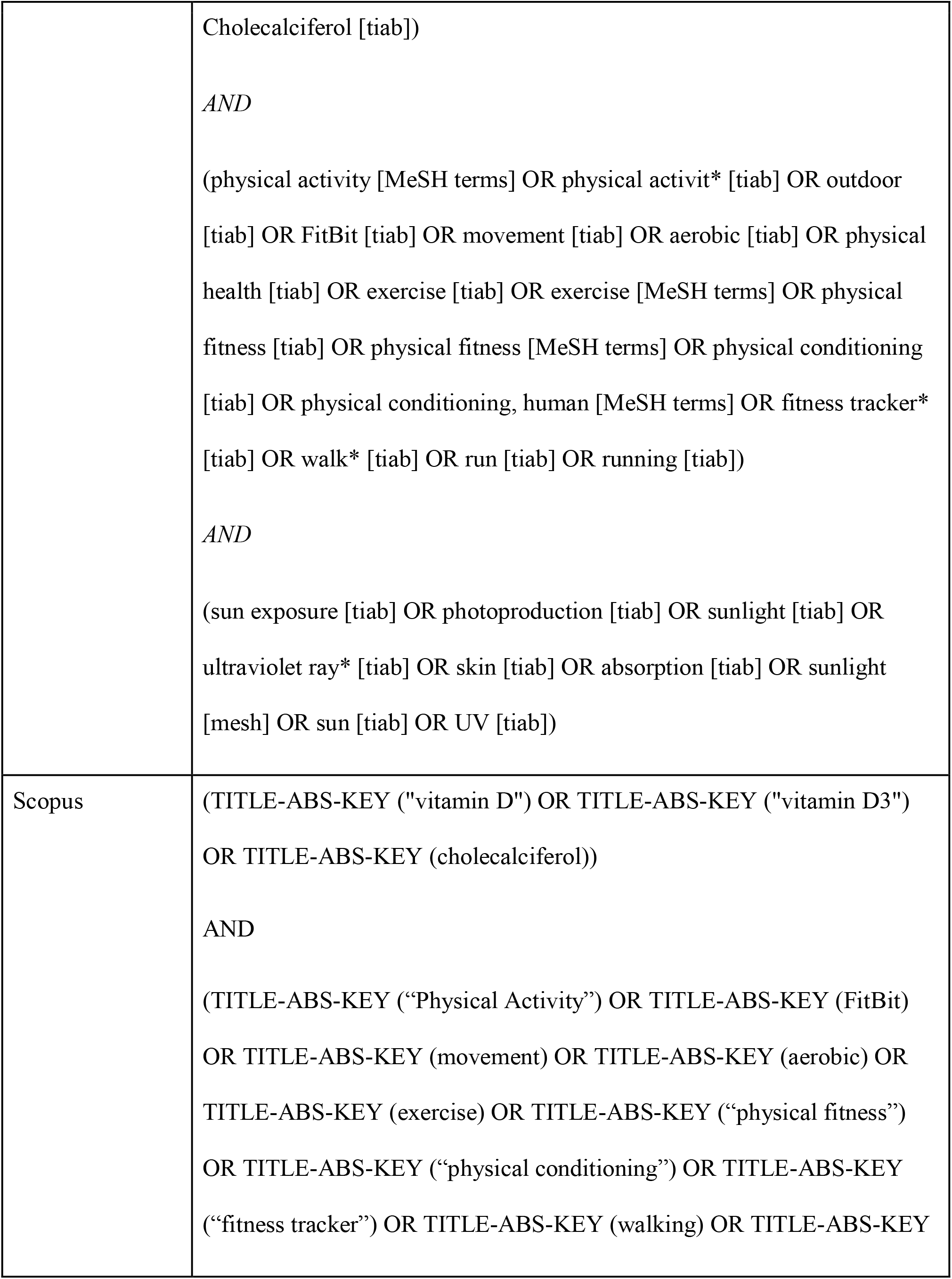
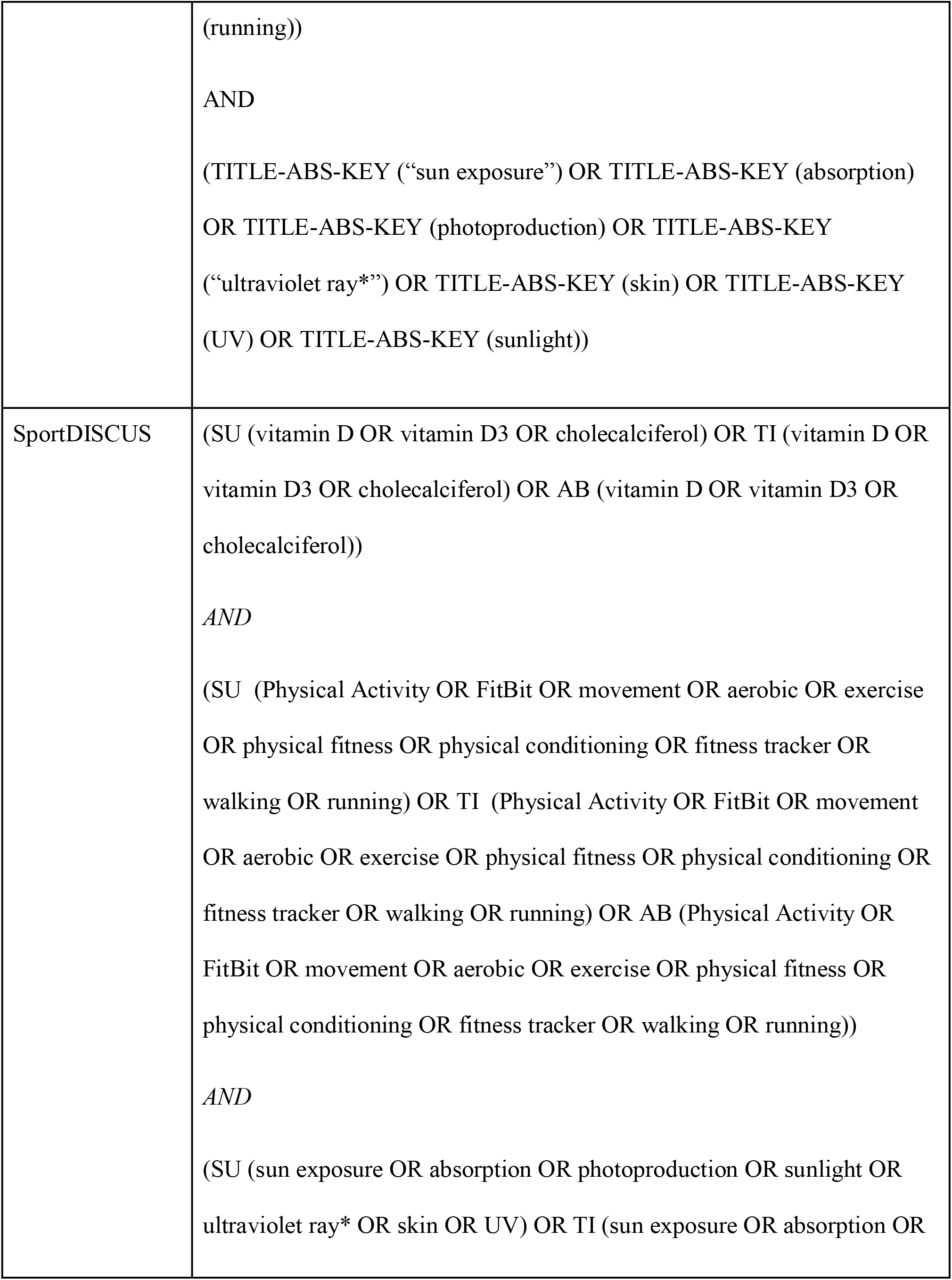
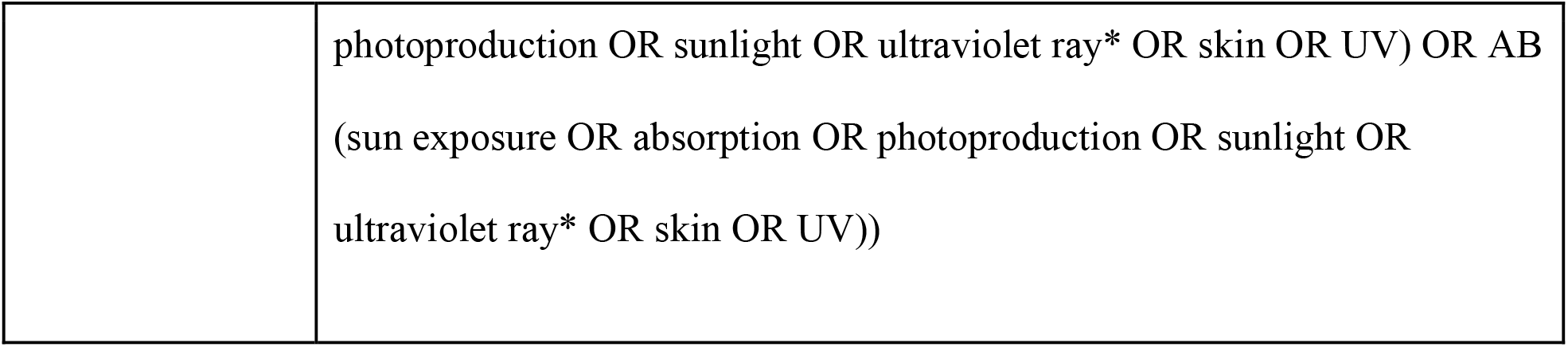
Database Search Strategies

### Inclusion and Exclusion Criteria

Consistent with Arksey and O’Malley’s process of conducting scoping reviews, the research team has developed specific inclusion and exclusion criteria to ensure we are collecting relevant and appropriate publications [1]. As discussed, the researchers are pulling literature from five electronic databases and, at initial search, have identified 2270 potentially relevant documents. To ensure that our search is strategic, we have identified the following inclusion criteria:

1. Published in English
2. Peer-reviewed publication
3. Considers the role of photoproduced vitD in disease prevention/health promotion
4. Has identified any form of PA in the objectives, methods, results, discussions, or conclusion section(s) of the publication

Based on conversations with our reference librarian and the research team, the following exclusion criteria were identified:

1. Published in any language other than English
2. Register of clinical trials without available results (i.e. notices of studies in progress)
3. Informal publications (e.g. blog posts, non-referred publications)

### Stage Three: Study Selection

The third stage of the Arksey and O’Malley process entails the elimination of the studies irrelevant to the central research question [1]. The research team will create a set of more detailed inclusion and exclusion criteria, related to the source of vitD, the comparison of variables, and the outcomes on the populations being studied. These criteria will be applied to the determine the relevance of studies, first solely to the titles and abstracts of selected studies and then to the full text articles. From there, the extracted data will be synthesized and interpreted.

### Determining Relevance and Quality of Literature

The research team will conduct a three-phase process of reviewing publications for inclusion in this review. First, a title and abstract screening will be conducted to determine relevance given the questions guiding this study. In this first phase, two researchers will review each title and abstract to determine relevance. If there is not consensus among these two researchers, a senior researcher will provide a final determination. Second, the researchers will conduct a full-text review of each publication. In this phase, two researchers will be required to review each publication to determine whether it aligns with our specific inclusion criteria following the population, intervention, comparison, and outcome (PICO) guidelines [9]; again, a senior researcher will serve as a tiebreaker if necessary. If a study is excluded at full-text review, the rationale will be provided for publication in the PRISMA flow diagram. Finally, the third step will include data extraction of relevant information for thematic analysis. The extraction form guiding this final step of the scoping review is available in Table 2.

**Table 2:**
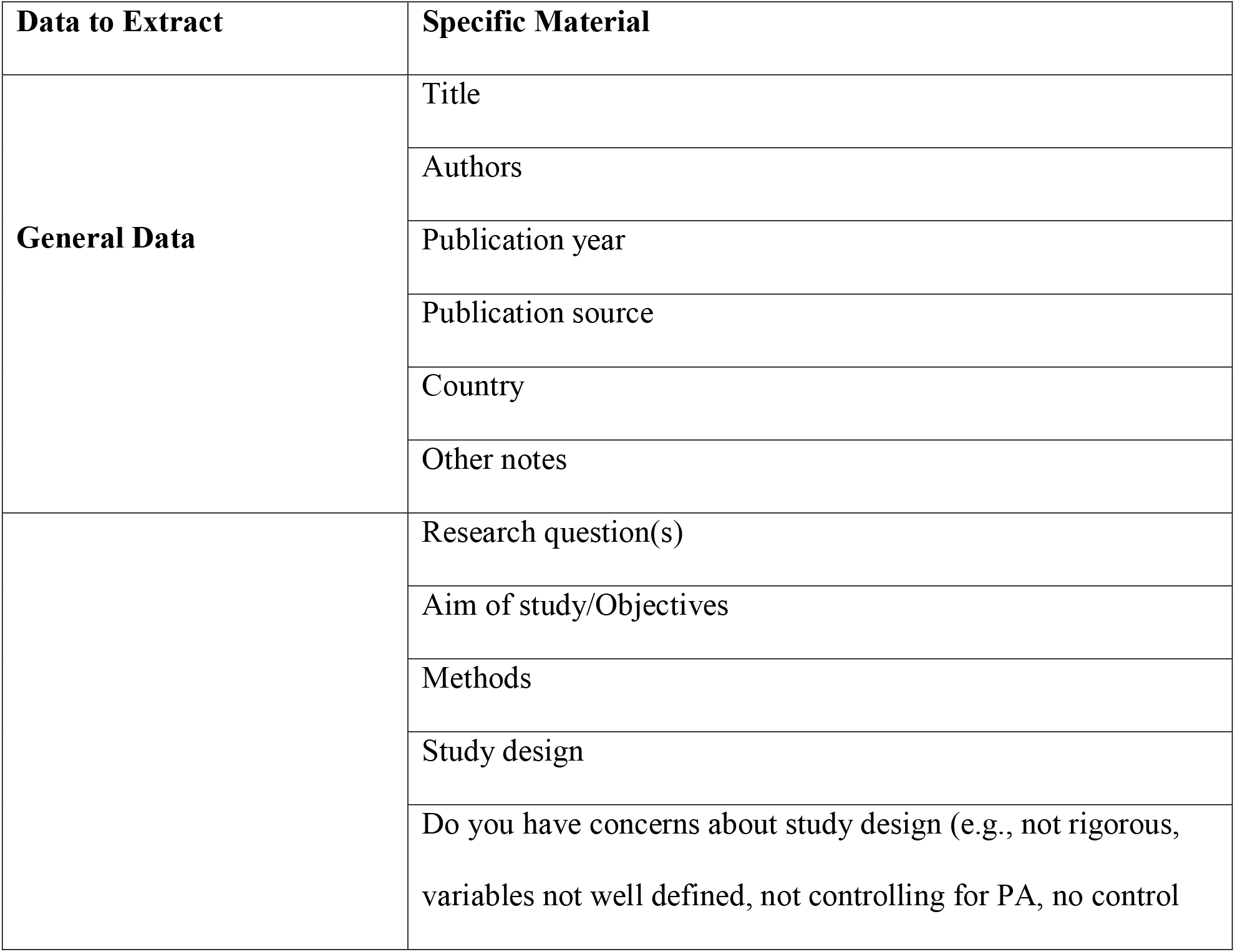
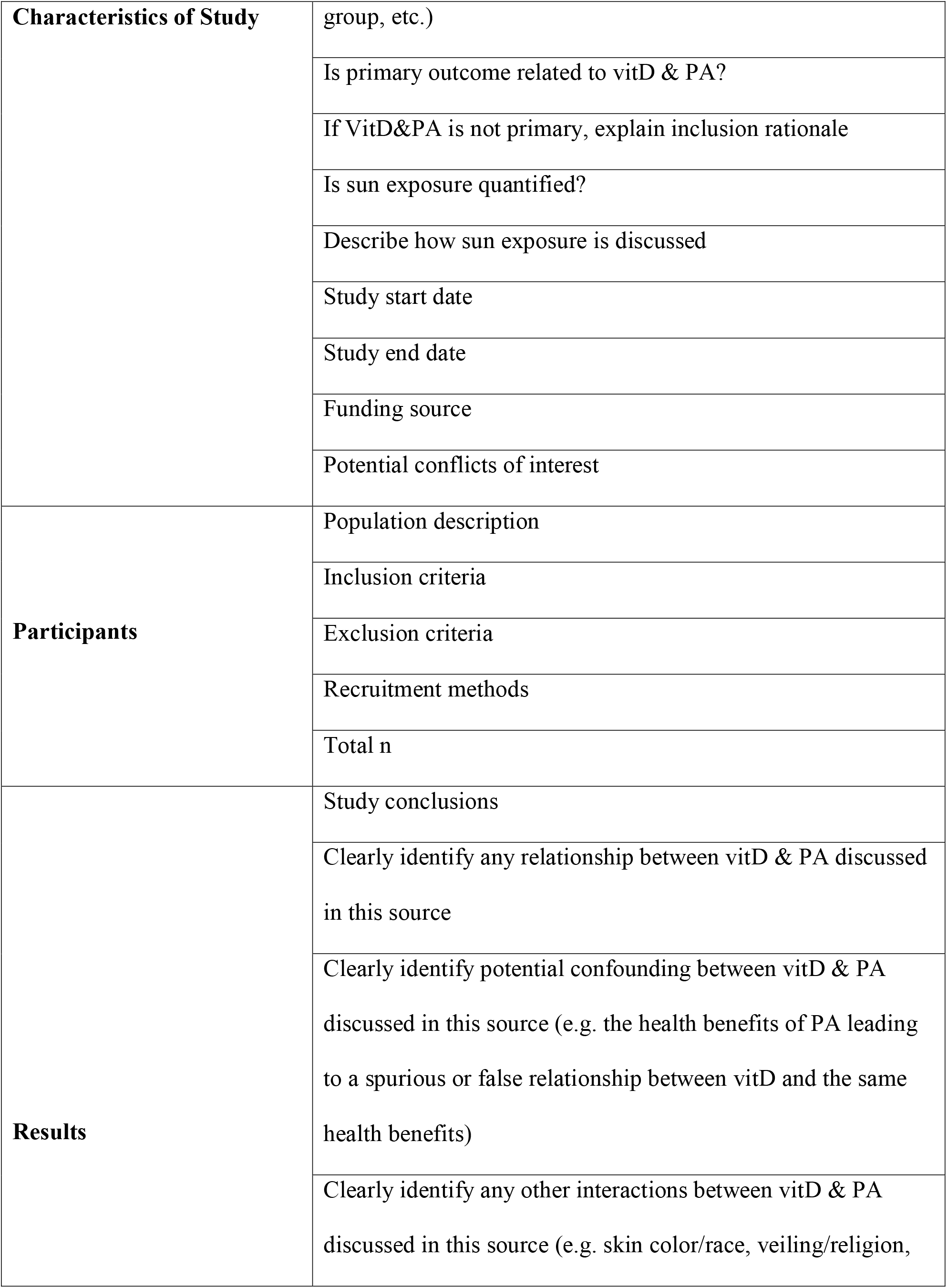
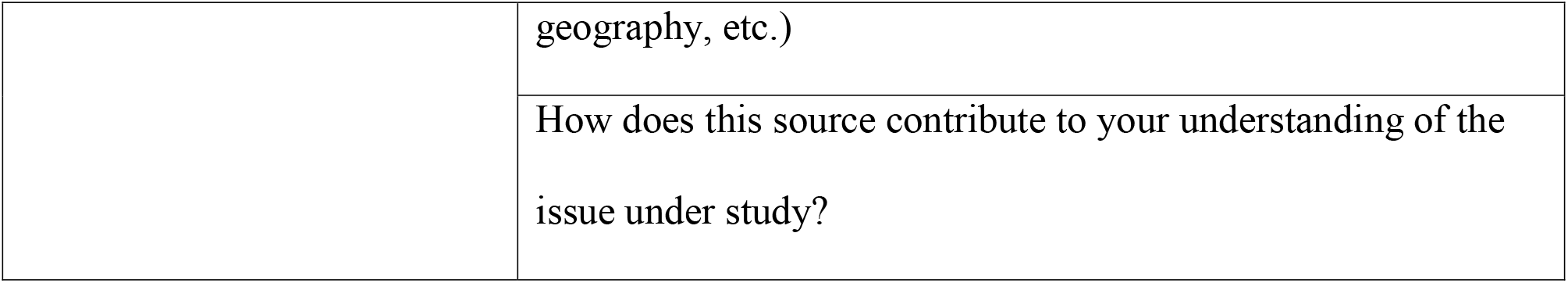
Covidence Data Extraction Form

### Stage Four: Charting the Data

The research team will use the data extraction form in the Covidence database, which provides a structured and auditable process to identify aspects of each study and ultimately to determine the relevance and/or limitations to our study. A summary of the specific material included in the form is provided in Table 2. This allows a uniform approach to all studies that are reviewed. From there, the findings of the extracted qualitative data will be synthesized, integrated, and interpreted into a thematic analysis process.

### Stage Five: Collecting, Summarizing, and Reporting the Data

This scoping review aims to identify and index existing evidence for confounding between vitD status and PA revealing a particular gap in the relationship between vitD and PA. The resulting information will be coded to provide an overview of how vitD markers in current research correlate with PA in addition to other measured health status indicators to map the relationship between these health measures and the potential for these relationships to be spurious. The resulting information will refine how the researchers analyze variables related to vitD and PA in the All of Us Research Database to better understand how PA impacts the relationship with vitD biomarkers and health outcomes [11].

### Limitations

Because qualitative data is often specific to each study based on its variables and population, the data is sometimes not generalizable. Therefore, it can be difficult to apply a uniform thematic analysis process to all studies. As such, there will be a set of limitations presented in this study. We anticipate that some articles will not specify details of the variables studied, such as the vitD source or the type and location of PA, while others may have possible confounding or influencing variables, including but not limited to skin color/race, veiling/religion, and geography. Additionally, some studies may possibly have conflicts of interest involving author affiliations or funding sources, and we hope to be able to assess this in this scoping review. The research team will work to identify and account for as many of these factors as possible.

## Conclusion

VitD is a key nutrient for bone maintenance and extra-skeletal effects have recently been elucidated, primarily in immune homeostasis and surveillance. The primary driver of vitD status is exposure to UV-B radiation, which is also associated with outdoor PA. When studying the health outcomes associated with vitD status, it is unclear what effect outdoor PA may have on these relationships and if they are, in fact, spurious, as the effect is, at least in part, due to fitness or other effects from outdoor PA. This scoping review will be the first step in exploring the relationship between vitD and PA to determine the strength and magnitude of potential confounders. Ultimately, we would like to estimate the bias produced by this relationship to be able to reassess previous findings and determine the true effect of vitD alone by isolating the effect of outdoor PA.

## Data Availability

No datasets were generated or analysed during the current study. All relevant data from this study will be made available upon study completion.

## Acknowledgments

The authors would like to acknowledge Thomas Harrod, Associate Director of Reference, Instruction, and Access at the George Washington University’s Himmelfarb Health Sciences Library for his generous guidance and support in developing the search strategies for this scoping review.

## Author Contributors

Conceptualization, PGC and LAF; Methodology, PGC, AMB, SHV, LAF; Writing – Original Draft Preparation, PGC, AMB, SHV; Writing – Review & Editing, LAF; Supervision, LAF.

## Institutional Review Board Statement

Not applicable.

